# Determinants of COVID-19 vaccine uptake in the Netherlands: an ecological analysis

**DOI:** 10.1101/2023.02.01.23284949

**Authors:** Lisanne J.E. Labuschagne, Naomi Smorenburg, Jan van de Kassteele, Ben Bom, Anne de Weerdt, Hester E. de Melker, Susan J.M. Hahné

## Abstract

**Background:** While overall COVID-19 vaccine uptake is high in the Netherlands, it lags behind in certain subpopulations.

**Aim:** We aimed to identify determinants associated with COVID-19 vaccine uptake at neighbourhood level to inform the strategy to improve uptake and guide research into barriers for vaccination. We focused on those aged 50 years and older, since they are at highest risk of severe disease.

**Methods:** We performed an ecological study using national vaccination register and socio-demographic data at neighbourhood level. Using univariate and multivariable generalized additive models we examined the (potentially non-linear) effect of each determinant on uptake.

**Results:** In those over 50 years of age, a higher proportion of individuals with a non-Western migration background and higher voting proportions for right-wing Christian and conservative political parties were at neighbourhood level univariately associated with lower COVID-19 vaccine uptake. In contrast, higher socioeconomic status and higher voting proportions for right-wing liberal, progressive liberal and Christian middle political parties were associated with higher uptake. Multivariable results differed from univariate results in that a higher voting proportion for progressive left-wing political parties was also associated with higher uptake. In addition, with regard to migration background only a Turkish background remained significant.

**Conclusion:** We identified determinants associated with COVID-19 vaccine uptake at neighbourhood level and observed heterogeneity between different subpopulations. Since the goal of the vaccination campaign is not only to reduce suffering and death by improving the average uptake, but also to reduce health inequity, it is important to focus on these hard-to-reach populations.

## Introduction

The COVID-19 vaccination campaign in the Netherlands started 6 January 2021. The first groups targeted were employees in direct COVID-19 patient care, general practitioners, residents of long term care facilities and other persons living in an institution. In the context of vaccine shortage, the vaccination strategy was to offer vaccination from old to young [1]. By the end of June 2022, the coverage for at least one dose of COVID-19 vaccination for individuals aged 12 years or over was approximately 83%, while 82% was fully vaccinated [2]. Although the overall vaccination coverage was high and the coverage among individuals aged 50 years and older was above 90%, vaccination coverage of younger age groups lagged behind [1-2]. In addition, the uptake was lower in the four biggest cities and in so-called ‘Bible Belt’ municipalities where relatively many orthodox reformed individuals reside, who are known to refuse vaccination more often [1]. To improve our understanding of COVID-19 vaccination behaviour, we performed an ecological study at neighbourhood level. This allowed studying a wide variety of potential determinants which are not yet available for studies at an individual level. The aim of the study was to identify determinants at neighbourhood level that are (independently) associated with COVID-19 vaccine uptake in the Netherlands, in order to improve the strategy to increase uptake and guide research into barriers for vaccination.

## Methods

### Vaccine uptake

Vaccine uptake was calculated with data from the COVID vaccine Information- and Monitoring System (CIMS). CIMS is a nationwide register including all individuals who are registered in the national population register of the Netherlands. COVID-19 vaccinations are included for vaccinated individuals who have consented for this information to be registered in CIMS until April 12^th^ 2022. Approximately 93% of those vaccinated by municipal health services gave consent [3]. Thus, individuals for whom no vaccinations are registered in CIMS are either unvaccinated or did not give consent for their vaccination to be registered. ‘Vaccine uptake’ was defined as having received at least one COVID-19 vaccine. It was not possible to examine coverage (i.e. a completed primary series of COVID-19 vaccination), since we did not have data on SARS-CoV-2 infections, which rendered one dose to be sufficient. Vaccine uptake per neighbourhood was stratified by age group (12-49 and 50+ years). We focused our analyses on the 50+ age group, as high uptake is particularly important in those considering that COVID-19 is more severe at older age.

### Determinants

Potential determinants at neighbourhood level were extracted from the publicly available data of Statistics Netherlands (CBS), which included information regarding migration background, socioeconomic status and urbanisation [4]. A neighbourhood is defined as a part of a municipality dominated by a given type of land use or buildings, for instance: industrial area, residential area with high-rise or low-rise buildings. Neighbourhoods are subdivided into smaller neighbourhood areas with a homogenous socio-economic structure or planning (definition CBS) [4]. Results from the National Elections in March 2021 per voting location were available from the Open State Foundation. These results were then translated to voting proportions per neighbourhood (see Supplementary material 1 with a detailed list of political parties) [5]. Distance to nearest vaccination location was calculated as the distance from the centroid of the neighbourhood to the nearest vaccination facility, not including mobile vaccination facilities. Locations of the facilities in use in July 2021, at the peak of the large-scale vaccination campaign, were used. Finally, at the municipality level we obtained information about HPV vaccine uptake in 2020 among girls aged 14 years who were invited for HPV vaccination within the Dutch national immunisation program (NIP) [6].

### Statistical analyses

To examine possible associations between COVID-19 vaccine uptake and each determinant at neighbourhood level, we performed univariate and multivariable generalized additive models with a binomial outcome using a logit-link function. In this way we examined the (potentially non-linear) effect of each determinant on COVID-19 vaccine uptake, while correcting for effects of other determinants. More specifically, we used a quasi-binomial model. This model has an extra parameter that attempts to describe additional variance in the data that cannot be explained by a binomial distribution alone, e.g. caused by neighbourhood specific random effects.

As a first step, we carried out univariate analyses. We subsequently added specific (groups of) variables to the multivariable model to gain more insight into the interrelationships between factors, moving from distal to more proximate factors [7]. First, a model was estimated which only included migration background. In the second model, socioeconomic status was added. Finally, the third model also included urbanisation, distance to nearest vaccination location and voting proportions. Each determinant was included as a penalized spline to model potential non-linear effects. Highly (right) skewed determinants were first transformed to a more uniform or normal scale, e.g. by log- or square root transformation. Effects were presented graphically as odds-ratios of the likelihood to be vaccinated relative to the global average of the specific determinant. In addition, Spearman rank correlations between all determinants, including HPV vaccine uptake, were calculated. All analyses were done using the mgcv package in R [8].

## Results

By April 12^th^ 2022, the national overall COVID-19 vaccine uptake as registered in CIMS was 87.6% among individuals aged 50 years and older and 72.0% among individuals aged 12-49 years. Results of those aged 50 years and older were considered as main results given the highest risk of severe COVID-19. Analyses of the age group 12-49 years can be found in Supplementary Material 3-4.

### Determinants associated with COVID-19 vaccine uptake

The baseline characteristics of all (populated) 3243 neighbourhoods in the Netherlands are presented in Table 1. An overview of the direction of the associations and the significance of the determinants in each model for the population aged 50 years and older along with the explained variance (multivariable models) is presented in Table 2. It must be noted, however, that the direction of the association is based on a subjective interpretation of the graphs. We present these graphical results of the final model (model 3) in Figure 1 and those of the univariate analyses, model 1 and 2 in Supplementary Material 2.

**Table 1.**
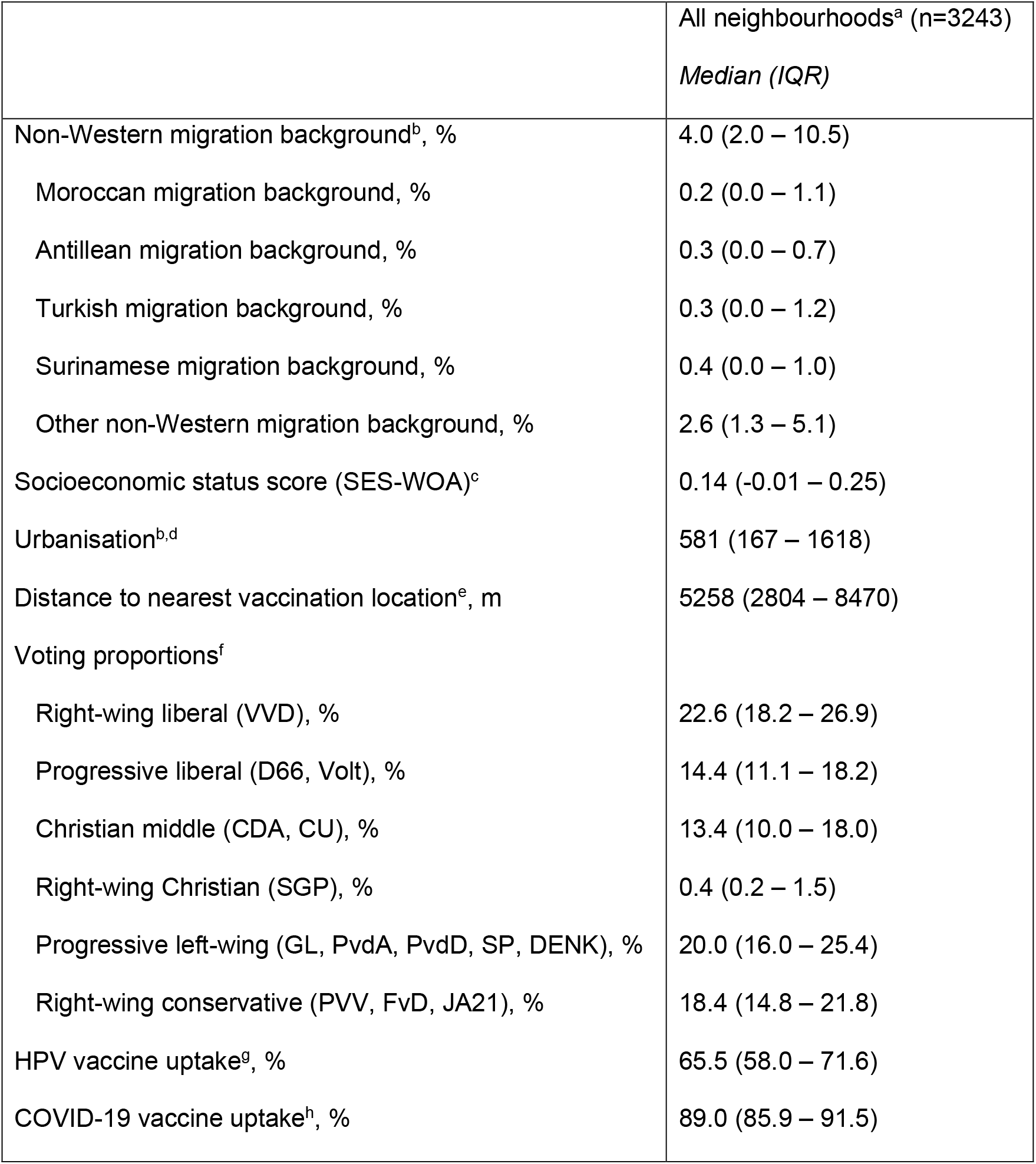
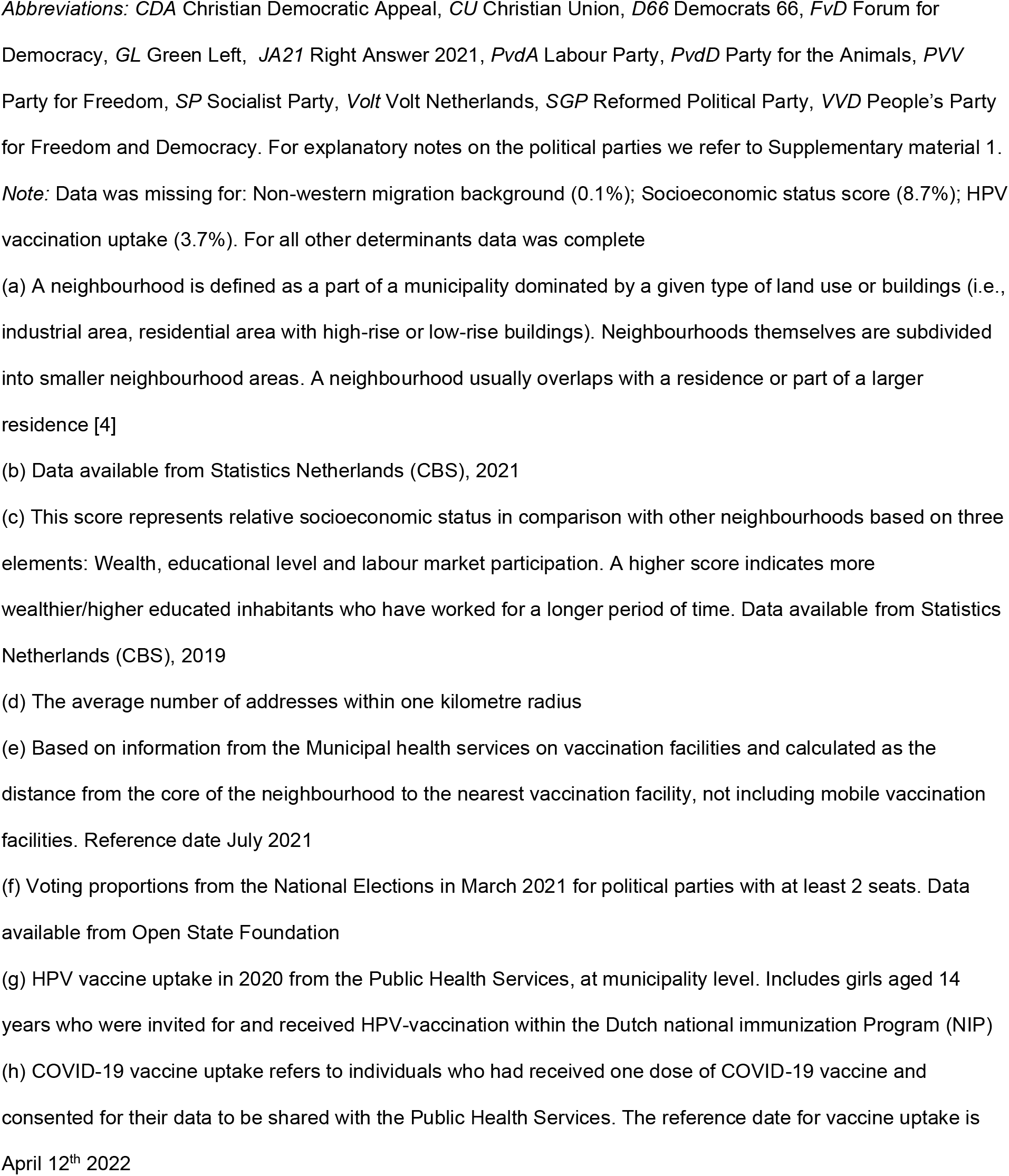
Baseline characteristics of included neighbourhoods, the Netherlands

|  | All neighbourhoods<sup>a</sup> (n=3243)<br>*Median (IQR)* |
| --- | --- |
| Non-Western migration background<sup>b</sup>, % | 4.0 (2.0 – 10.5) |
| Moroccan migration background, % | 0.2 (0.0 – 1.1) |
| Antillean migration background, % | 0.3 (0.0 – 0.7) |
| Turkish migration background, % | 0.3 (0.0 – 1.2) |
| Surinamese migration background, % | 0.4 (0.0 – 1.0) |
| Other non-Western migration background, % | 2.6 (1.3 – 5.1) |
| Socioeconomic status score (SES-WOA)<sup>c</sup> | 0.14 (-0.01 – 0.25) |
| Urbanisation<sup>b,d</sup> | 581 (167 – 1618) |
| Distance to nearest vaccination location<sup>e</sup>, m | 5258 (2804 – 8470) |
| Voting proportions<sup>f</sup> |  |
| Right-wing liberal (VVD), % | 22.6 (18.2 – 26.9) |
| Progressive liberal (D66, Volt), % | 14.4 (11.1 – 18.2) |
| Christian middle (CDA, CU), % | 13.4 (10.0 – 18.0) |
| Right-wing Christian (SGP), % | 0.4 (0.2 – 1.5) |
| Progressive left-wing (GL, PvdA, PvdD, SP, DENK), % | 20.0 (16.0 – 25.4) |
| Right-wing conservative (PVV, FvD, JA21), % | 18.4 (14.8 – 21.8) |
| HPV vaccine uptake<sup>g</sup>, % | 65.5 (58.0 – 71.6) |
| COVID-19 vaccine uptake<sup>h</sup>, % | 89.0 (85.9 – 91.5) |

*Abbreviations:* CDA Christian Democratic Appeal, CU Christian Union, D66 Democrats 66, FvD Forum for Democracy, GL Green Left, JA21 Right Answer 2021, PvdA Labour Party, PvdD Party for the Animals, PVV Party for Freedom, SP Socialist Party, Volt Volt Netherlands, SGP Reformed Political Party, VVD People's Party for Freedom and Democracy. For explanatory notes on the political parties we refer to Supplementary material 1.
*Note:* Data was missing for: Non-western migration background (0.1%); Socioeconomic status score (8.7%); HPV vaccination uptake (3.7%). For all other determinants data was complete
(a) A neighbourhood is defined as a part of a municipality dominated by a given type of land use or buildings (i.e., industrial area, residential area with high-rise or low-rise buildings). Neighbourhoods themselves are subdivided into smaller neighbourhood areas. A neighbourhood usually overlaps with a residence or part of a larger residence [4]
(b) Data available from Statistics Netherlands (CBS), 2021
(c) This score represents relative socioeconomic status in comparison with other neighbourhoods based on three elements: Wealth, educational level and labour market participation. A higher score indicates more wealthier/higher educated inhabitants who have worked for a longer period of time. Data available from Statistics Netherlands (CBS), 2019
(d) The average number of addresses within one kilometre radius
(e) Based on information from the Municipal health services on vaccination facilities and calculated as the distance from the core of the neighbourhood to the nearest vaccination facility, not including mobile vaccination facilities. Reference date July 2021
(f) Voting proportions from the National Elections in March 2021 for political parties with at least 2 seats. Data available from Open State Foundation
(g) HPV vaccine uptake in 2020 from the Public Health Services, at municipality level. Includes girls aged 14 years who were invited for and received HPV-vaccination within the Dutch national immunization Program (NIP)
(h) COVID-19 vaccine uptake refers to individuals who had received one dose of COVID-19 vaccine and consented for their data to be shared with the Public Health Services. The reference date for vaccine uptake is April 12<sup>th</sup> 2022

**Table 2.** Results of univariate and multivariable analyses of the associations at neighbourhood level between potential determinants and COVID-19 vaccine uptake among individuals of 50 years and older.

|  | Univariate |  | Multivariable |  |  |  |  |  |
| --- | --- | --- | --- | --- | --- | --- | --- | --- |
| | | | Model 1<br>($R^2 = 17.1\%$) | | Model 2<br>($R^2 = 20.7\%$) | | Model 3<br>($R^2 = 41.7\%$) | |
| | direction of<br>association | $p$ | direction of<br>association | $p$ | direction of<br>association | $p$ | direction of<br>association | $p$ |
| Non-Western migration<br>background: |  |  |  |  |  |  |  |  |
| Moroccan | – | < .001 | – | < .001 | – | < .001 | – | 0.487 |
| Antillean | – | < .001 | – | < .001 | 0 | 1.000 | 0 | 0.731 |
| Turkish | – | < .001 | – | 0.001 | – | 0.006 | – | 0.026 |
| Surinamese | – | < .001 | – | 0.012 | – | 0.017 | – | 0.171 |
| Other | – | < .001 | – | < .001 | – | 0.020 | – | 0.137 |
| Higher socioeconomic status | + | < .001 |  |  | + | < .001 | + | < .001 |
| Higher degree of urbanisation | ~ | < .001 |  |  |  |  | + | < .001 |
| Larger distance to nearest vaccination location | ~ | < .001 |  |  |  |  | 0 | 0.185 |
| Voting proportions |  |  |  |  |  |  |  |  |
| Right-wing liberal | + | < .001 |  |  |  |  | + | 0.001 |
| Progressive liberal | + | < .001 |  |  |  |  | + | < .001 |
| Christian middle | + | < .001 |  |  |  |  | + | < .001 |
| Right-wing Christian | – | < .001 |  |  |  |  | – | < .001 |
| Progressive left-wing | ~ | < .001 |  |  |  |  | + | 0.002 |
| Right-wing conservative | – | < .001 |  |  |  |  | – | < .001 |
| HPV vaccine uptake | + | <.001 |  |  |  |  |  |  |

Note: All associations are at neighbourhood level. For all models the direction of association (positive (+), negative,(–), no association (0) and mixed (~)) and significance of determinants are presented. For multivariable models 1, 2 and 3, the explained variance is also included. The covariate included in model 1 was: migration background. Covariates included in model 2 were: migration background and socioeconomic status. Covariates included in model 3 were: migration background, socioeconomic status, urbanisation, distance to nearest vaccination location and voting proportions.

**Figure 1.**
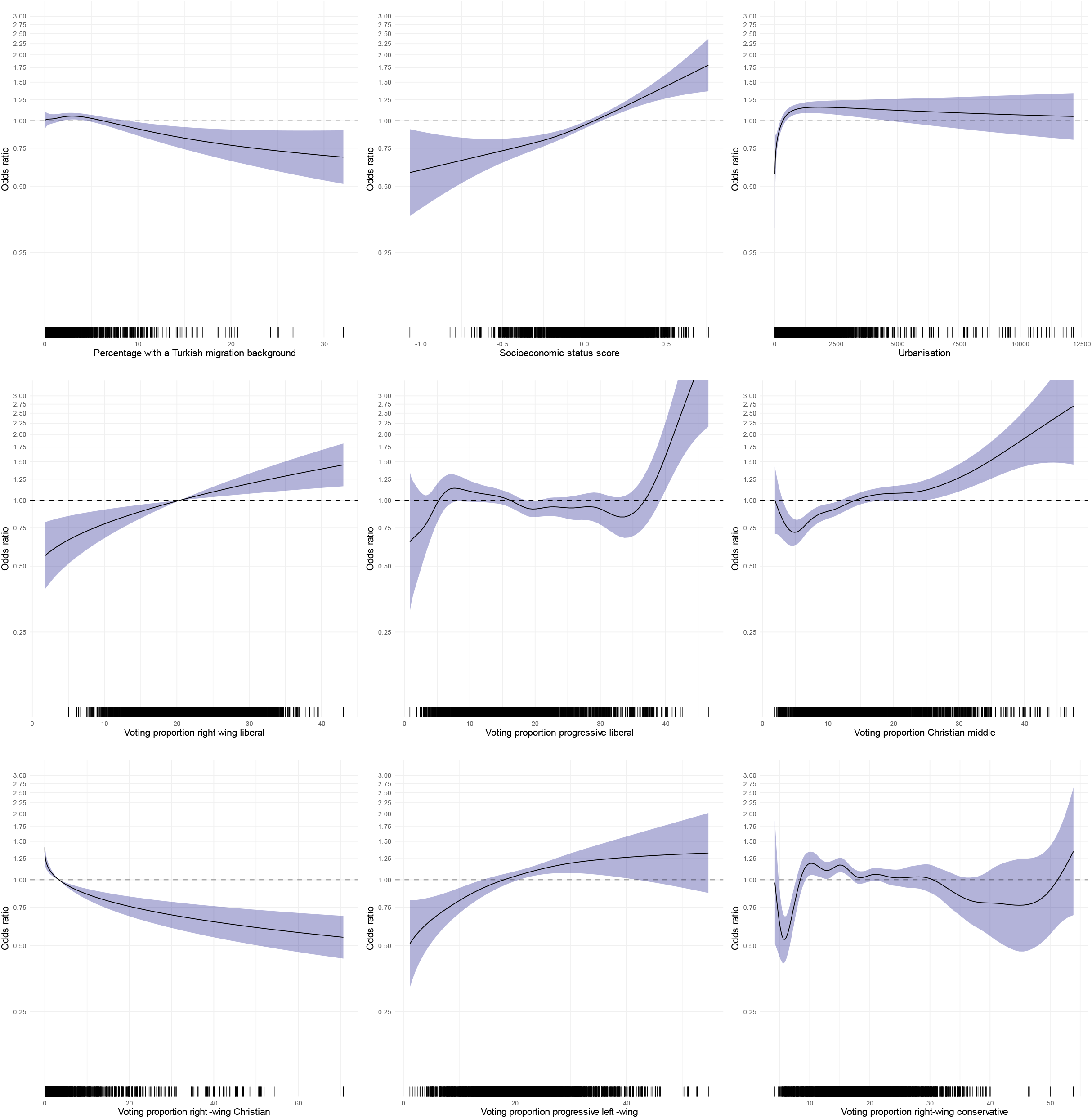
Multivariable binomial logistic regression analyses of the association at neighbourhood level between COVID-19 vaccine uptake and Turkish migration background, socioeconomic status score, urbanisation, and voting proportions for right-wing liberal, progressive liberal, Christian middle, right-wing Christian, progressive left-wing and right-wing conservative political parties (Model 3).

All determinants were significantly associated with COVID-19 vaccine uptake among individuals of 50 years of age and older in univariate analyses (Table 2). With a rising percentage of individuals with any type of non-Western migration background, the odds for vaccine uptake for COVID-19 at the neighbourhood level decreased (Table 2 and Supplementary Figure 2.1). Higher voting proportions for right-wing Christian and right-wing conservative political parties were also associated with a lower uptake. On the other hand, higher socioeconomic status, higher voting proportions for right-wing liberal, progressive liberal and Christian middle political parties and higher HPV vaccine uptake were univariately associated with higher COVID-19 vaccine uptake. For a higher degree of urbanisation, a higher distance to nearest vaccination location and higher voting proportions for progressive left-wing political parties, the association with COVID-19 vaccine uptake was mixed, meaning that the association was positive or negative depending on the prevalence of the determinant.

In multivariable analyses, the percentages of individuals with all types of non-Western migration background were significantly negatively associated with COVID-19 vaccine uptake in Model 1 (see Table 2 and Supplementary Figure S2.2). When socioeconomic status was added to the model (Model 2), the association for ‘Antillean’ migration background was no longer significant. In the final model (Model 3), including all potential determinants, for migration background only the association with ‘Turkish’ migration background remained significant.

Higher socioeconomic status was significantly positively associated with COVID-19 vaccine uptake in multivariable analyses (Model 2) and remained significant when other determinants were added in Model 3 (see Figure 1 Supplementary Figure S2.2). Higher voting proportions for right-wing liberal, progressive liberal, Christian middle and progressive left-wing political parties were also positively associated with COVID-19 vaccine uptake (Model 3). For higher degree of urbanisation, the association was also significant and positive but only up to a certain level of urbanisation, after which the association stabilised (Model 3, see Figure 1). Distance to nearest vaccination location was not significantly associated with COVID-19 vaccine uptake (Model 3).

### Correlations between determinants

Figure 2 presents the Spearman rank correlations between all potential determinants on neighbourhood level and COVID-19 vaccine uptake. Some determinants were highly correlated. For example, all non-Western migration backgrounds were highly positively correlated with urbanisation, and voting proportions for progressive liberal and progressive left-wing political parties and negatively correlated with socioeconomic status, Christian middle political parties and distance to nearest vaccination location. Furthermore, correlations between all non-Western migration backgrounds were very strong, which might explain why only Turkish migration background remained significant in the final model. In addition, urbanisation was significantly negatively correlated with socioeconomic status and distance to nearest vaccination location. This suggests that in more densely populated areas people did not have to travel far to be vaccinated, but these also happened to be places where socioeconomic status was lower and a relatively larger group of migrants lived. Finally, HPV vaccine uptake was highly correlated with COVID-19 vaccine uptake.

**Figure 2.**
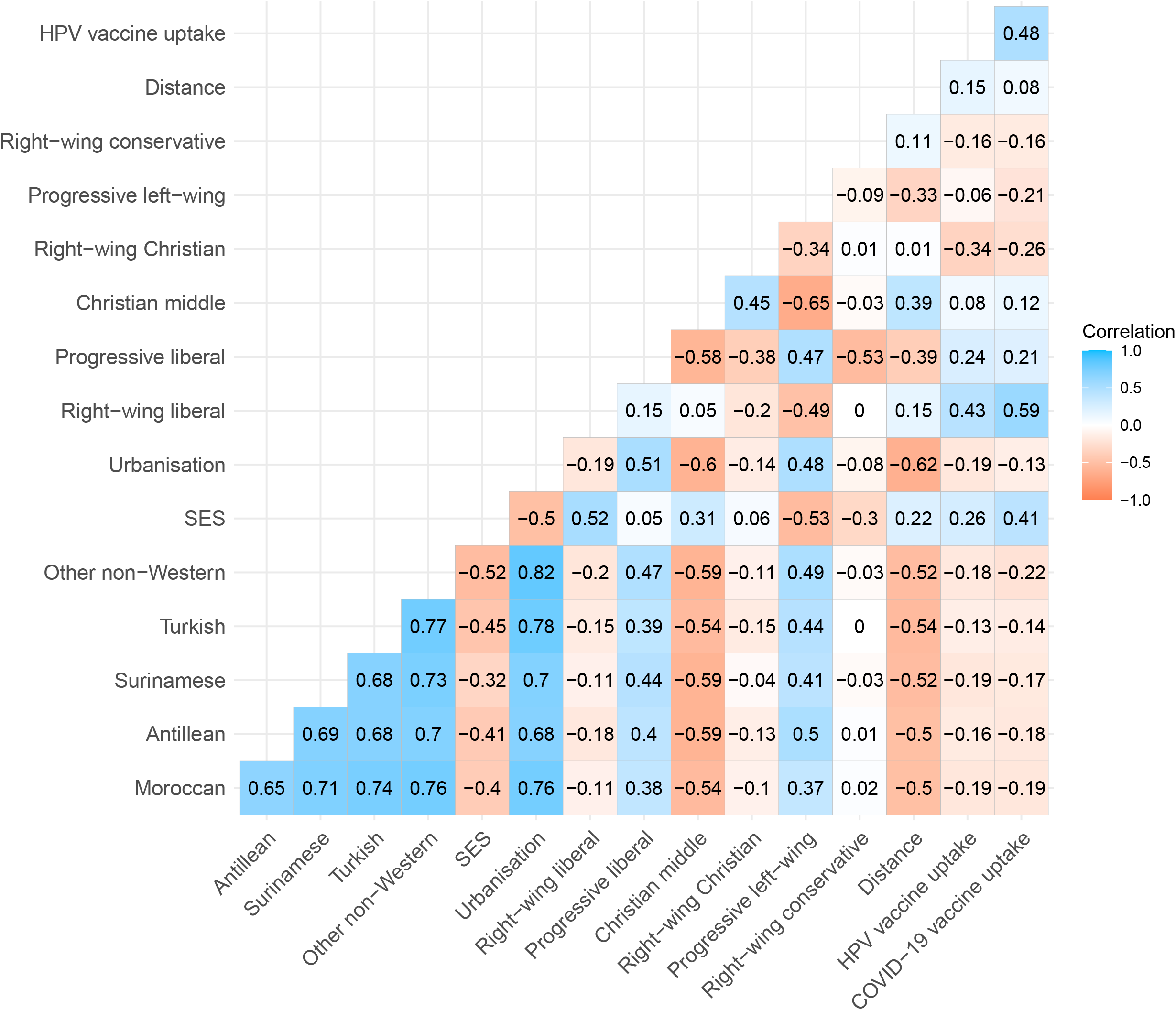
Correlations between all potential determinants, HPV vaccine uptake and COVID-19 vaccine uptake among individuals of 50 years and older at neighbourhood level.

### Differences with age group 12-49 years

Results for the age group 12-49 years were mostly similar to those aged 50 years and older (see Supplementary Material 3-4), but there are some differences worth mentioning. First of all, while the effects of most non-Western migration backgrounds became non-significant in the final model in those over 50 years of age, in the group aged 12-49 years these associations remained significant, except for Antillean migration background. The directions of the associations were very similar. With respect to voting proportions, the most striking difference was that right-wing liberal voting proportions were negatively associated with vaccine uptake in the final model, while this association was positive for those over 50 years of age. However, in univariate analyses the association was also positive for the younger age group. The association with progressive left-wing voting proportions was not significant. In the final model for those aged 12-49 years, substantially more variance was explained (*R*^2^ = 66.5%) compared to the model for the older age group (*R*^2^ = 41.7%).

## Discussion

While various studies reported on determinants for the intention to vaccinate, we were able to study determinants that were independently associated with actual vaccine uptake at neighbourhood level and identify possible population subgroups that might require more attention. In univariate analyses of the COVID-19 vaccine uptake among individuals of 50 years and older, we found that lower uptake was found in neighbourhoods with relatively many people with Moroccan, Antillean, Turkish, Surinamese or ‘other non-Western’ migration backgrounds and also in neighbourhoods with higher voting proportions for right-wing Christian and right-wing conservative political parties. In contrast, higher voting proportions for right-wing liberal, progressive liberal and Christian middle political parties and a higher socio-economic status were univariately associated with higher COVID-19 vaccine uptake. Our multivariable results showed that among individuals of 50 years and older independent negative determinants for vaccine uptake were a higher percentage of individuals with a Turkish migration background and higher percentages of voters for right-wing Christian and right-wing conservative political parties. Clear independent positive determinants were: a higher socioeconomic status and higher voting proportions for right-wing liberal, progressive liberal, Christian middle and progressive left-wing political parties.

These results are largely in line with small survey studies investigating vaccination willingness or hesitancy that have been performed in the Netherlands, indicating that individuals with a non-Western migration background and/or a lower socioeconomic status are less likely to be vaccinated against COVID-19 [9,10]. Our findings are also consistent with earlier studies in the Netherlands concerning other vaccination types. Parents’ country of birth, percentage of votes for the conservative Christian reformed party and low educational level have been associated with both lower HPV [6] and MenACWY-vaccine uptake [11]. In addition, in our study, HPV-vaccination background was strongly positively associated with COVID-19 vaccine uptake.

In international studies on willingness to vaccinate, similar findings were reported. In a large systematic review about factors that influence unwillingness or hesitancy to vaccinate against COVID-19 among older individuals, the likelihood of being unvaccinated was significantly higher in ethnic minority groups, or individuals with a low education or low income [12]. Multiple studies have confirmed that older individuals [13-15] and individuals with higher socioeconomic status [13,14, 16-18] were more likely to report the intention to be vaccinated against COVID-19. Being unemployed [19], having an ethnic minority status [20] and living in disadvantaged areas [21] were factors associated with lower willingness to be vaccinated against COVID-19. Our results are consistent with these determinants of willingness to vaccinate.

Results on urbanisation were more difficult to interpret. As the degree of urbanisation increased, the likelihood to be vaccinated against COVID-19 first increased. However, at a higher level this effect stabilised. Neighbourhoods with a higher degree of urbanisation thus had higher COVID-19 vaccine uptake compared to very unpopulated areas, but after a certain threshold, vaccine uptake did not differ. Distance to nearest vaccination location was not significant. It should be noted, however, that we were not able to include mobile vaccination locations. In addition, distance was highly correlated with urbanisation and non-Western migration background, which might have rendered it redundant in the multivariable analyses.

Results from the analyses among individuals aged 12-49 years were similar to the main results although uptake was lower in this group. Exceptions were, in the multivariable results, stronger significant negative associations for percentage of non-Western migration backgrounds, a non-significant association with voting proportion for progressive left-wing parties and vaccine uptake and a negative instead of a positive association with voting proportion for right-wing liberal parties. However, in univariate analyses the latter association was also positive for the younger age group. After correcting for other neighbourhood characteristics, different factors therefore seem to play a role in COVID-19 vaccine uptake in this younger age group compared to those aged 50 years and older.

The main strength of our study is that we were able to investigate COVID-19 vaccine uptake directly in contrast to previous studies that only concerned individuals’ willingness to be vaccinated before the vaccine was actually available. Our study also has some limitations. We performed ecological analyses at neighbourhood level, which requires the results to be interpreted with caution due to the problem of ecologic fallacy. In addition, since we only had data on individuals who had consented for their vaccination status to be shared with the Public Health Services, we were essentially investigating determinants for vaccine uptake *and* informed consent.

### Conclusion

Even though in the Netherlands overall COVID-19 vaccine uptake is high, we observed important heterogeneity between different subpopulations at neighbourhood level. Our results require further investigation and this study can therefore be considered as a first step to guide further research into what determinants might play a role in COVID-19 vaccine uptake and what population subgroups require more attention in vaccination campaigns. Further research on the role of the current determinants in COVID-19 vaccine uptake at an individual level and underlying reasons for not being vaccinated is recommended and underway. This is of key importance, since the goal of the vaccination programme is to not only prevent suffering and death by improving the average uptake, but also to reduce health inequity [22].

## Data Availability

All data produced in the present study are available in aggregated and anonymized form upon reasonable request to the authors.

## Contributions

LL, HdM, SH designed the study; NS, LL, BB, AdW generated the data; JvdK, NS, LL, AdW, performed analyses; SH, HdM supervised the study; LL, NS, HdM, SH wrote the manuscript.

