## Supplementary material for "Determinants of COVID-19 vaccine uptake in the Netherlands: an ecological analysis"

### Supplementary materials

This supplementary material is hosted by *Eurosurveillance* as supporting information alongside the article Determinants of COVID-19 vaccination uptake in the Netherlands: an ecological analysis, on behalf of the authors, who remain responsible for the accuracy and appropriateness of the content. The same standards for ethics, copyright, attributions and permissions as for the article apply. Supplements are not edited by *Eurosurveillance* and the journal is not responsible for the maintenance of any links or email addresses provided therein.

**Table S1.1. Political parties included in the in the Dutch House of Representatives in 2021**

| Abbreviation in Dutch | Party's name in English | Political movement / political spectrum |
|---|---|---|
| VVD | People's Party for Freedom and Democracy | Right-wing liberal party with more progressive positions in ethical matters |
| D66 | Democrats 66 | Reformist social-liberal party |
| Volt | Volt Netherlands | Social-liberal pro-Europeanism party |
| CDA | Christian Democratic Appeal | Christian-inspired party at the center of the political spectrum |
| CU | Christian Union | Christian party, with progressive positions in the social and ecological field and conservative positions on ethical issues |
| SGP | Reformed Political Party | Conservative Christian (Reformed) party that wants to conduct politics strictly according to Biblical standards |
| GL | Green Left | Progressive party which attaches great importance to sustainability |
| PvdA | Labor Party | Progressive, social-democratic party |
| PvdD | The Party for the Animals | Testimonial party with as main goals animal rights and animal welfare |
| SP | Socialist Party | Socialist, Eurosceptic party which has a strong local, action-oriented basis |
| DENK | Denk | Movement for migrants and a "tolerant and solidary society" |
| PVV | Party for Freedom | Populist party with both conservative, liberal "right" and "left" views |
| FvD | Forum for Democracy | Conservative, right-wing populist Eurosceptic political party |
| JA21 | Right Answer 2021 | Conservative right-wing liberal party, split from FvD |

Note: Political movements were grouped in the following manner: Right-wing liberal: VVD; Progressive liberal: D66, Volt; Christian middle: CDA, CU; Right-wing Christian: SGP; Progressive left-wing: GL, PvdA, PvdD, SP, DENK; Right-wing conservative: PVV, FvD, JA21.

*Figure S2.1.* **Univariate binomial logistic regression analyses of the association at neighbourhood level between COVID-19 vaccine uptake and Moroccan, Antillean, Turkish, Surinamese, and ‘ other non-Western’ migration background, socioeconomic status score, urbanisation, distance to nearest vaccination location, voting proportions for right-wing liberal, progressive liberal, Christian middle, right-wing Christian, progressive left-wing and right-wing conservative political parties and HPV vaccine uptake.**

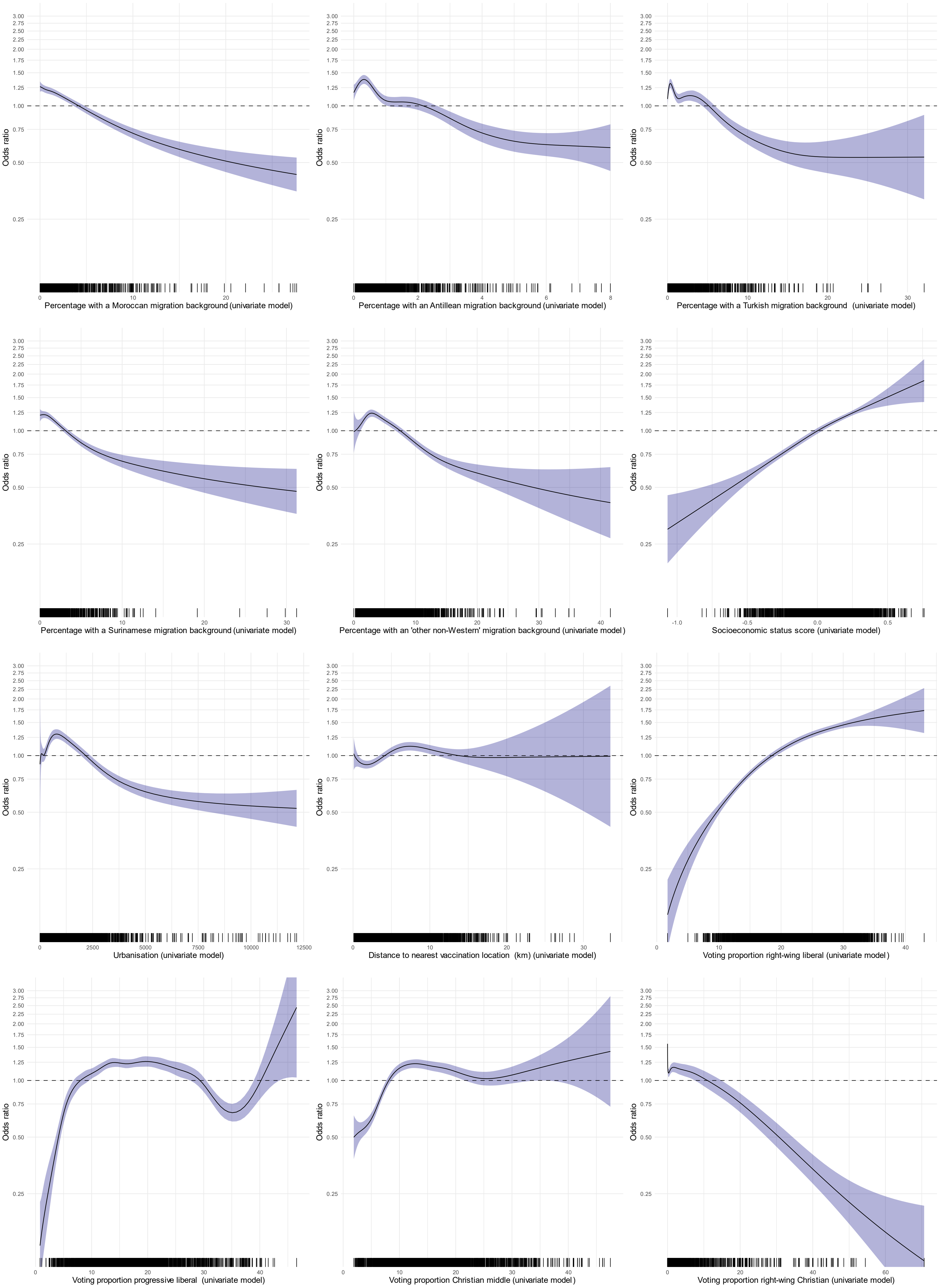
Odds ratio
3.00
2.75
2.50
2.25
2.00
1.75
1.50
1.25
1.00
0.75
0.50
0.25
Percentage with a Moroccan migration background (univariate model)
Percentage with an Antillean migration background (univariate model)
Percentage with a Turkish migration background (univariate model)
Percentage with a Surinamese migration background (univariate model)
Percentage with an 'other non-Western' migration background (univariate model)
Socioeconomic status score (univariate model)
Urbanisation (univariate model)
Distance to nearest vaccination location (km) (univariate model)
Voting proportion right-wing liberal (univariate model)
Voting proportion progressive liberal (univariate model)
Voting proportion Christian middle (univariate model)
Voting proportion right-wing Christian (univariate model)

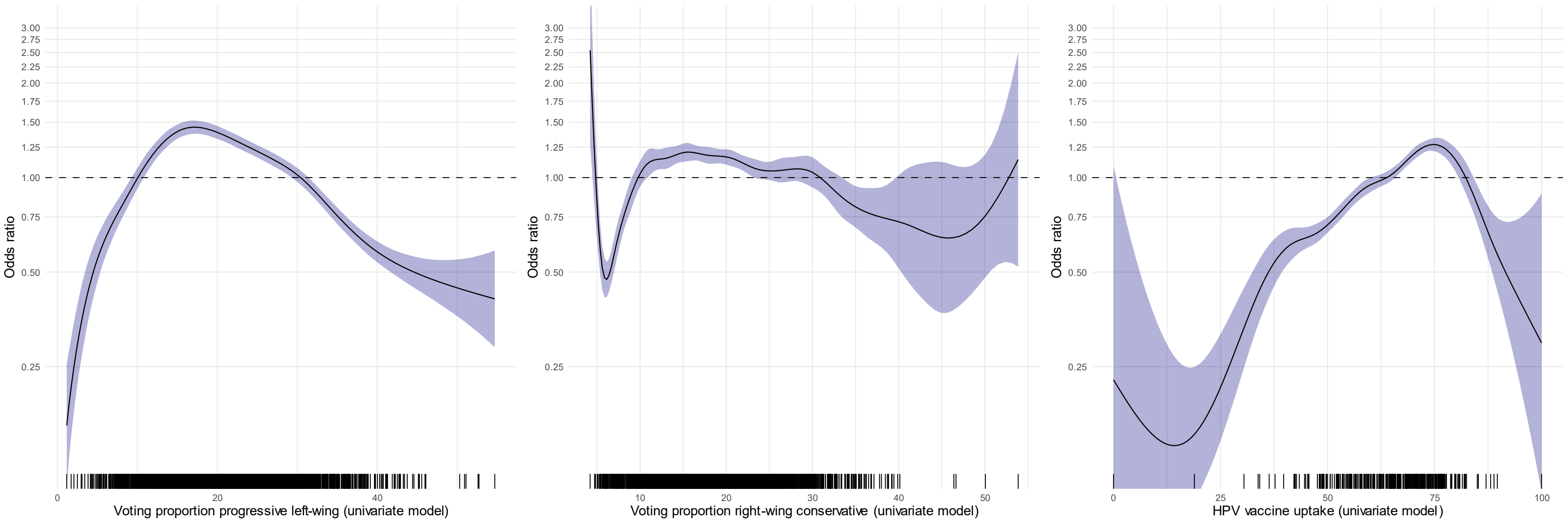
Odds ratio
3.00
2.75
2.50
2.25
2.00
1.75
1.50
1.25
1.00
0.75
0.50
0.25
0
20
40
Voting proportion progressive left-wing (univariate model)
10
20
30
40
50
Voting proportion right-wing conservative (univariate model)
0
25
50
75
100
HPV vaccine uptake (univariate model)

*Figure S2.2.* **Multivariable binomial logistic regression analyses of the association at neighbourhood level between COVID-19 vaccine uptake and Moroccan, Antillean, Turkish, Surinamese, and ‘ other non-Western’ migration background, socioeconomic status score and distance to nearest vaccination location (models 1, 2 and 3).**

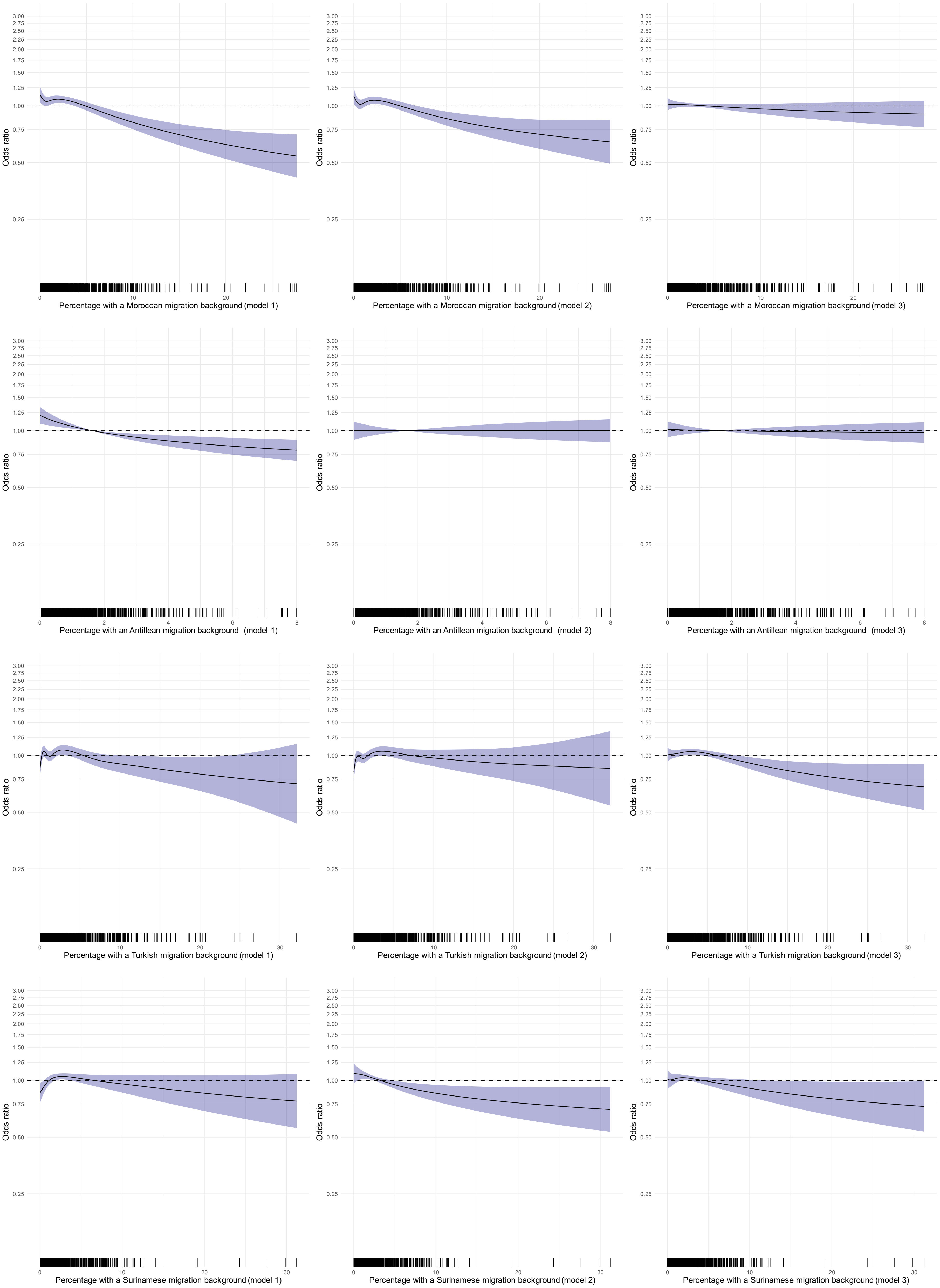

Odds ratio
3.00
2.75
2.50
2.25
2.00
1.75
1.50
1.25
1.00
0.75
0.50
0.25
Percentage with a Moroccan migration background (model 1)
Percentage with a Moroccan migration background (model 2)
Percentage with a Moroccan migration background (model 3)
Percentage with an Antillean migration background (model 1)
Percentage with an Antillean migration background (model 2)
Percentage with an Antillean migration background (model 3)
Percentage with a Turkish migration background (model 1)
Percentage with a Turkish migration background (model 2)
Percentage with a Turkish migration background (model 3)
Percentage with a Surinamese migration background (model 1)
Percentage with a Surinamese migration background (model 2)
Percentage with a Surinamese migration background (model 3)

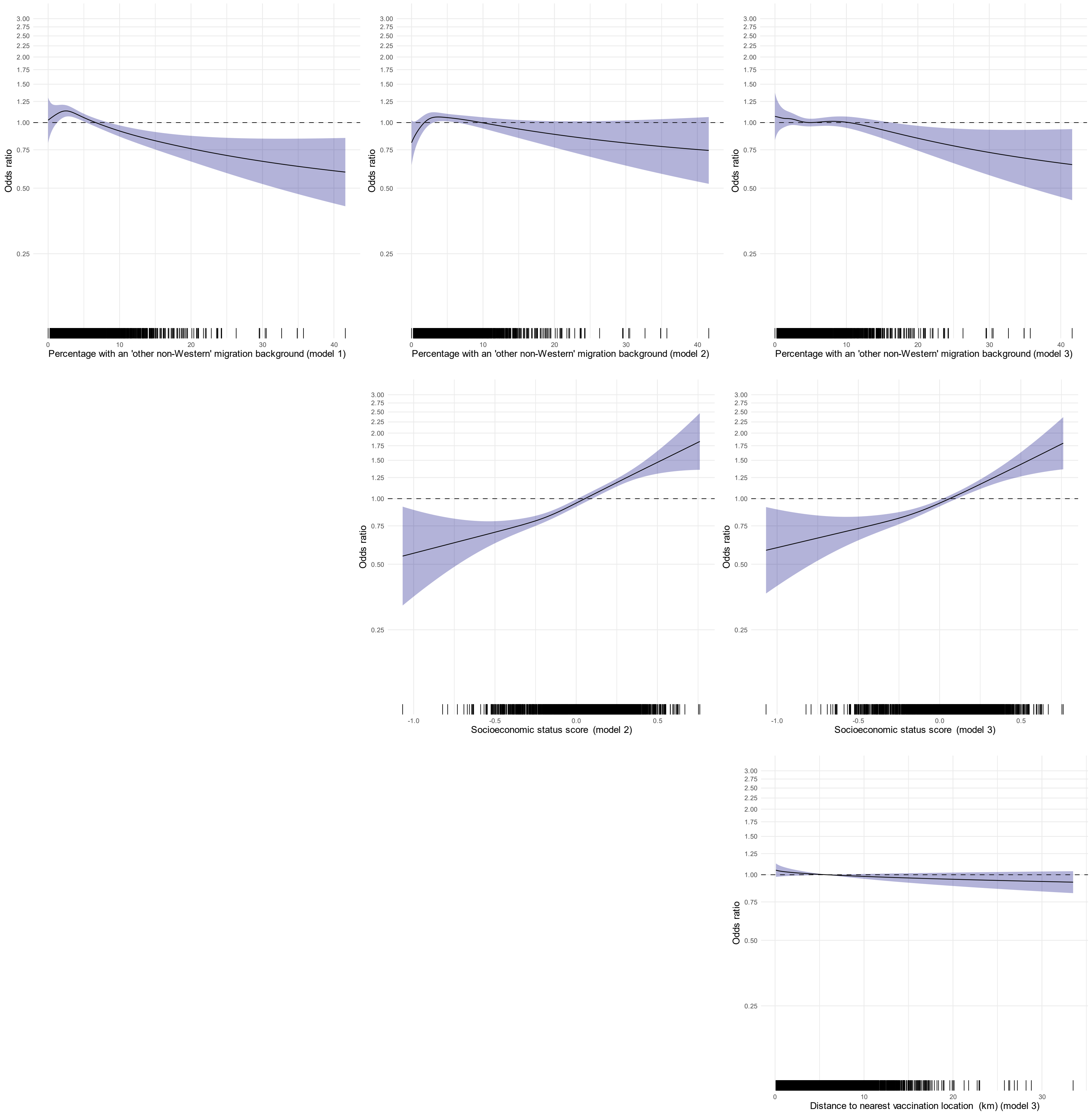
Odds ratio
3.00
2.75
2.50
2.25
2.00
1.75
1.50
1.25
1.00
0.75
0.50
0.25
0
10
20
30
40
Percentage with an 'other non-Western' migration background (model 1)
Percentage with an 'other non-Western' migration background (model 2)
Percentage with an 'other non-Western' migration background (model 3)
-1.0
-0.5
0.0
0.5
Socioeconomic status score (model 2)
Socioeconomic status score (model 3)
Distance to nearest vaccination location (km) (model 3)

**Table S3.1. Results of the univariate and multivariable analyses of the associations at neighbourhood level between potential determinants and COVID-19 vaccine uptake among individuals of 12-49 years.**

| | Univariate | | Multivariable | | | | | |
|---|---|---|---|---|---|---|---|---|
| | | | **Model 1** ($R^2$ = 36.2%) | | **Model 2** ($R^2$ = 42.6%) | | **Model 3** ($R^2$ = 66.5%) | |
| | direction of association | *p* | direction of association | *p* | direction of association | *p* | direction of association | *p* |
| Non-Western migration background: | | | | | | | | |
| Moroccan | − | < .001 | − | < .001 | − | < .001 | − | < .001 |
| Antillean | − | < .001 | − | < .001 | − | < .001 | 0 | 0.085 |
| Turkish | − | < .001 | ~ | < .001 | − | < .001 | − | < .001 |
| Surinamese | − | < .001 | − | < .001 | − | < .001 | − | < .001 |
| Other | − | < .001 | − | < .001 | − | 0.003 | − | < .001 |
| Higher socioeconomic status | + | < .001 | | | + | < .001 | + | < .001 |
| Higher degree of urbanisation | ~ | < .001 | | | | | + | < .001 |
| Larger distance to nearest vaccination location | ~ | < .001 | | | | | 0 | 0.313 |
| Voting proportions | | | | | | | | |
| Right-wing liberal | + | < .001 | | | | | − | < .001 |
| Progressive liberal | + | < .001 | | | | | + | < .001 |
| Christian middle | + | < .001 | | | | | + | 0.018 |
| Right-wing Christian | − | < .001 | | | | | − | < .001 |
| Progressive left-wing | ~ | < .001 | | | | | ~ | 0.181 |

| | | | | | | | | |
|---|---|---|---|---|---|---|---|---|
| Right-wing conservative | − | < .001 | | | | | − | < .001 |
| HPV vaccine uptake | + | <.001 | | | | | | |

Note: All associations are at neighbourhood level. For all models, the direction of association (positive (+), negative,(−), no association (0) and mixed (~)) and significance of determinants are presented. For multivariable models 1, 2 and 3, the explained variance is also included. The covariate included in model 1 was: migration background. Covariates included in model 2 were: migration background and socioeconomic status. Covariates included in model 3 were: migration background, socioeconomic status. urbanisation, distance to nearest vaccination location and voting proportions.

*Figure S3.2.* **Multivariable binomial logistic regression analyses of the association at neighbourhood level between COVID-19 vaccine uptake and Moroccan, Turkish, Surinamese, and 'other non-Western' migration background, socioeconomic status score, urbanisation, and voting proportions for right-wing liberal, progressive liberal, Christian middle, right-wing Christian and right-wing conservative political parties (final model) (age group 12-49 years).**

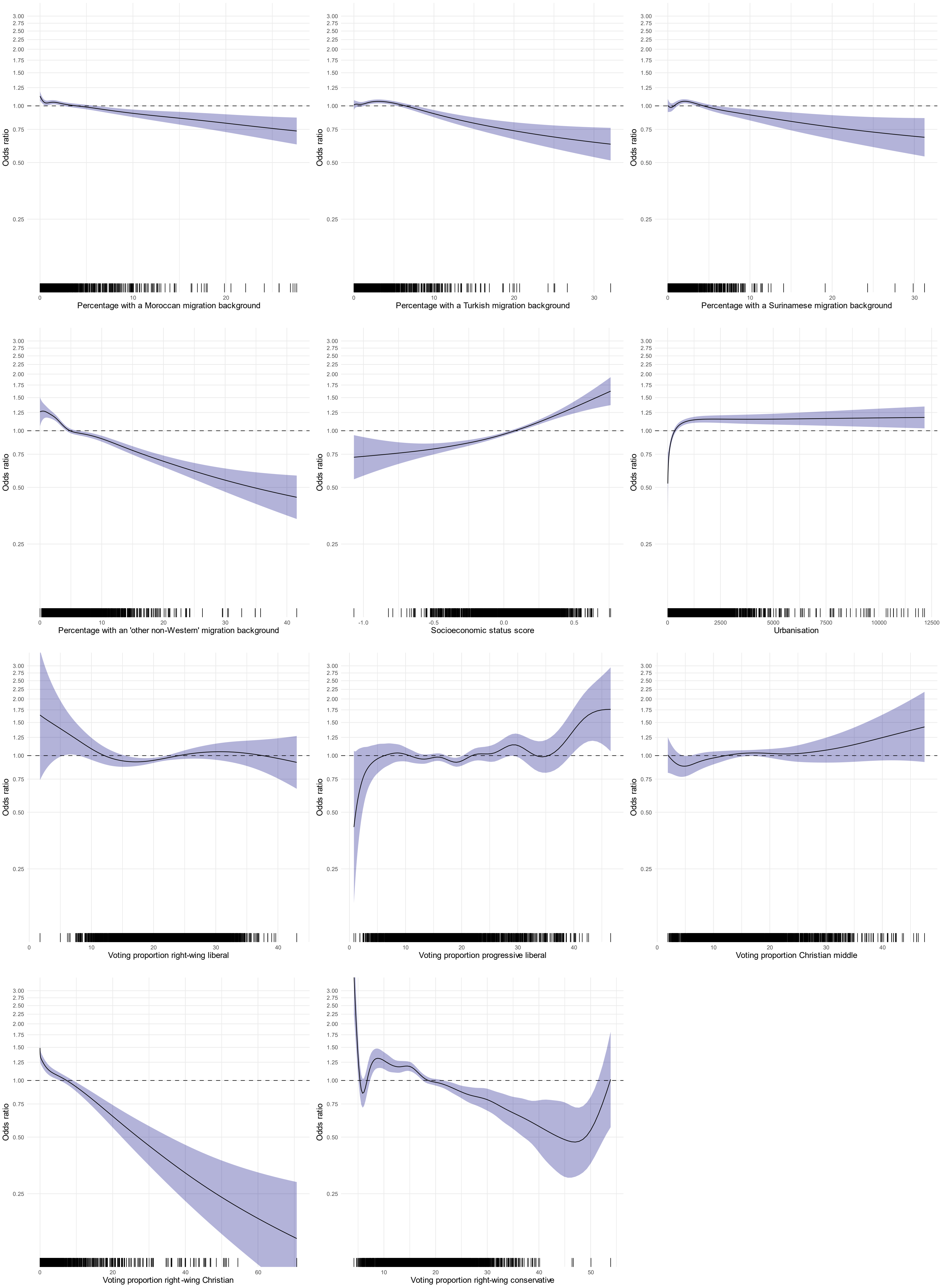
Odds ratio
3.00
2.75
2.50
2.25
2.00
1.75
1.50
1.25
1.00
0.75
0.50
0.25
Percentage with a Moroccan migration background
Percentage with a Turkish migration background
Percentage with a Surinamese migration background
Percentage with an 'other non-Western' migration background
Socioeconomic status score
Urbanisation
Voting proportion right-wing liberal
Voting proportion progressive liberal
Voting proportion Christian middle
Voting proportion right -wing Christian
Voting proportion right-wing conservative

*Figure S3.3.* **Correlations between all potential determinants, HPV vaccine uptake and COVID-19 vaccine uptake at neighbourhood level (age group 12-49 years).**

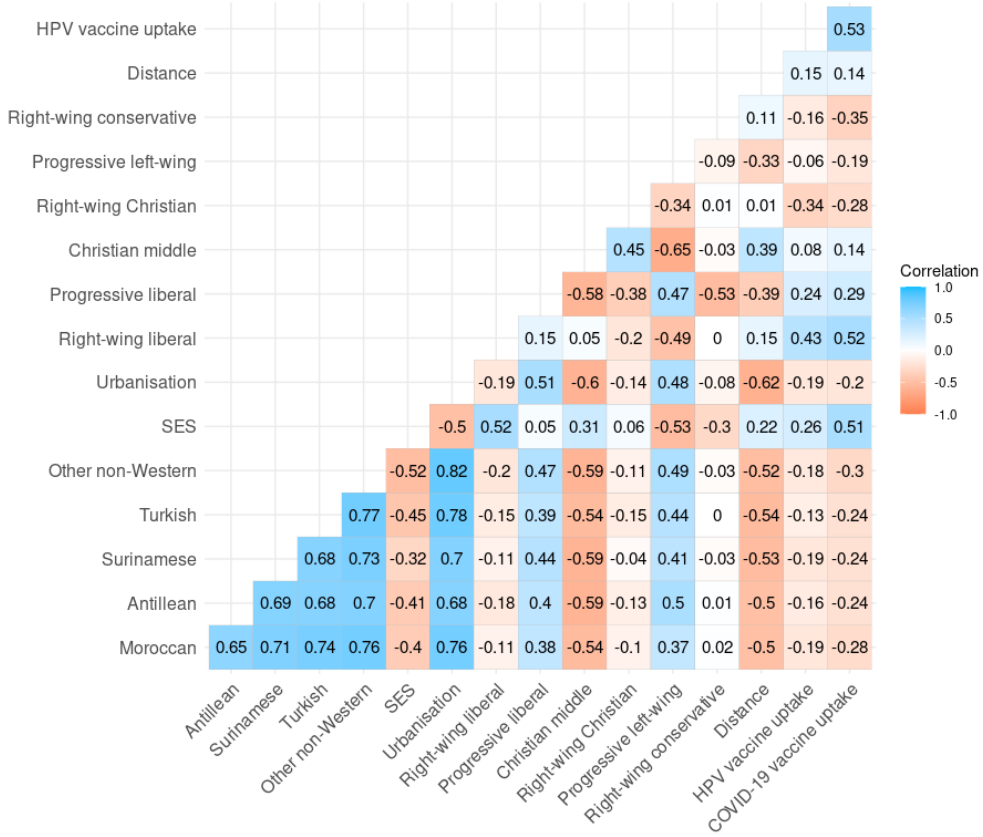

*Figure S4.1.* **Univariate binomial logistic regression analyses of the association at neighbourhood level between COVID-19 vaccine uptake and Moroccan, Turkish, Surinamese, and ‵ other non-Western′ migration background, socioeconomic status score, urbanisation, distance to nearest vaccination location, voting proportions for right-wing liberal, progressive liberal, Christian middle, right-wing Christian, progressive left-wing and right-wing conservative political parties and HPV vaccine uptake (age group 12-49 years).**

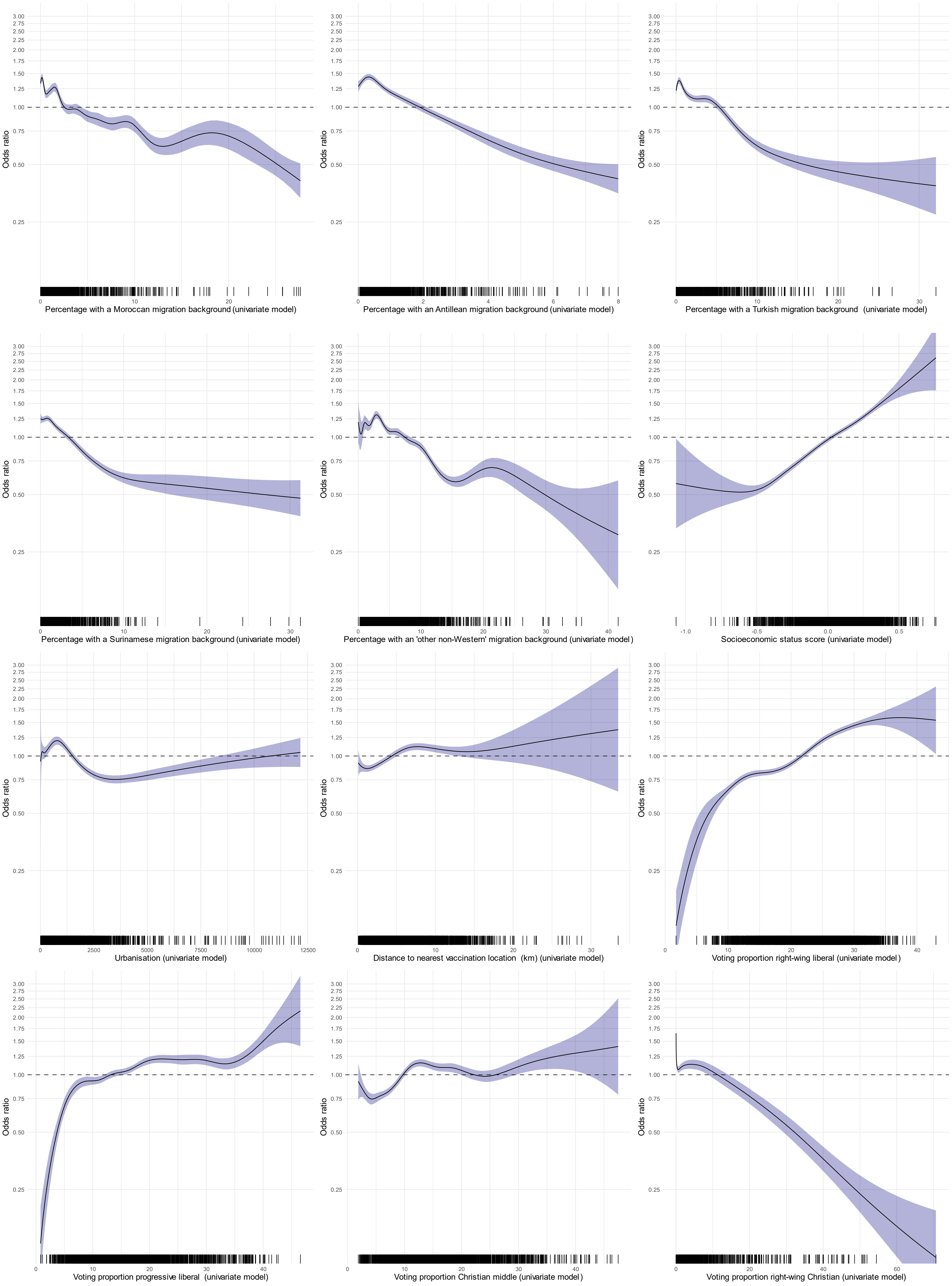
Odds ratio
3.00
2.75
2.50
2.25
2.00
1.75
1.50
1.25
1.00
0.75
0.50
0.25
Percentage with a Moroccan migration background (univariate model)
Percentage with an Antillean migration background (univariate model)
Percentage with a Turkish migration background (univariate model)
Percentage with a Surinamese migration background (univariate model)
Percentage with an 'other non-Western' migration background (univariate model)
Socioeconomic status score (univariate model)
Urbanisation (univariate model)
Distance to nearest vaccination location (km) (univariate model)
Voting proportion right-wing liberal (univariate model)
Voting proportion progressive liberal (univariate model)
Voting proportion Christian middle (univariate model)
Voting proportion right-wing Christian (univariate model)

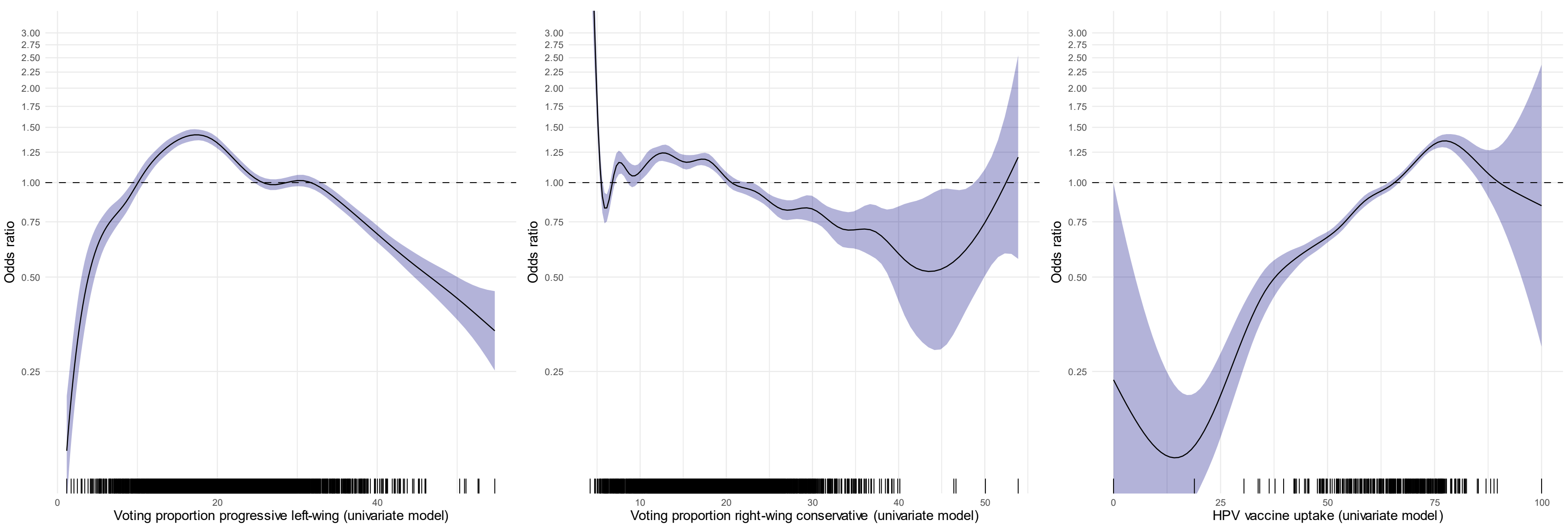
Odds ratio
3.00
2.75
2.50
2.25
2.00
1.75
1.50
1.25
1.00
0.75
0.50
0.25
0
20
40
Voting proportion progressive left-wing (univariate model)
10
20
30
40
50
Voting proportion right-wing conservative (univariate model)
0
25
50
75
100
HPV vaccine uptake (univariate model)

*Figure S4.2.* **Multivariable binomial logistic regression analyses of the association at neighbourhood level between COVID-19 vaccine uptake and Moroccan, Antillean, Turkish, Surinamese, and ‘ other non-Western’ migration background, socioeconomic status score, distance to nearest vaccination location and voting proportion for progressive left-wing political parties (models 1, 2 and 3) (age group 12-49 years).**

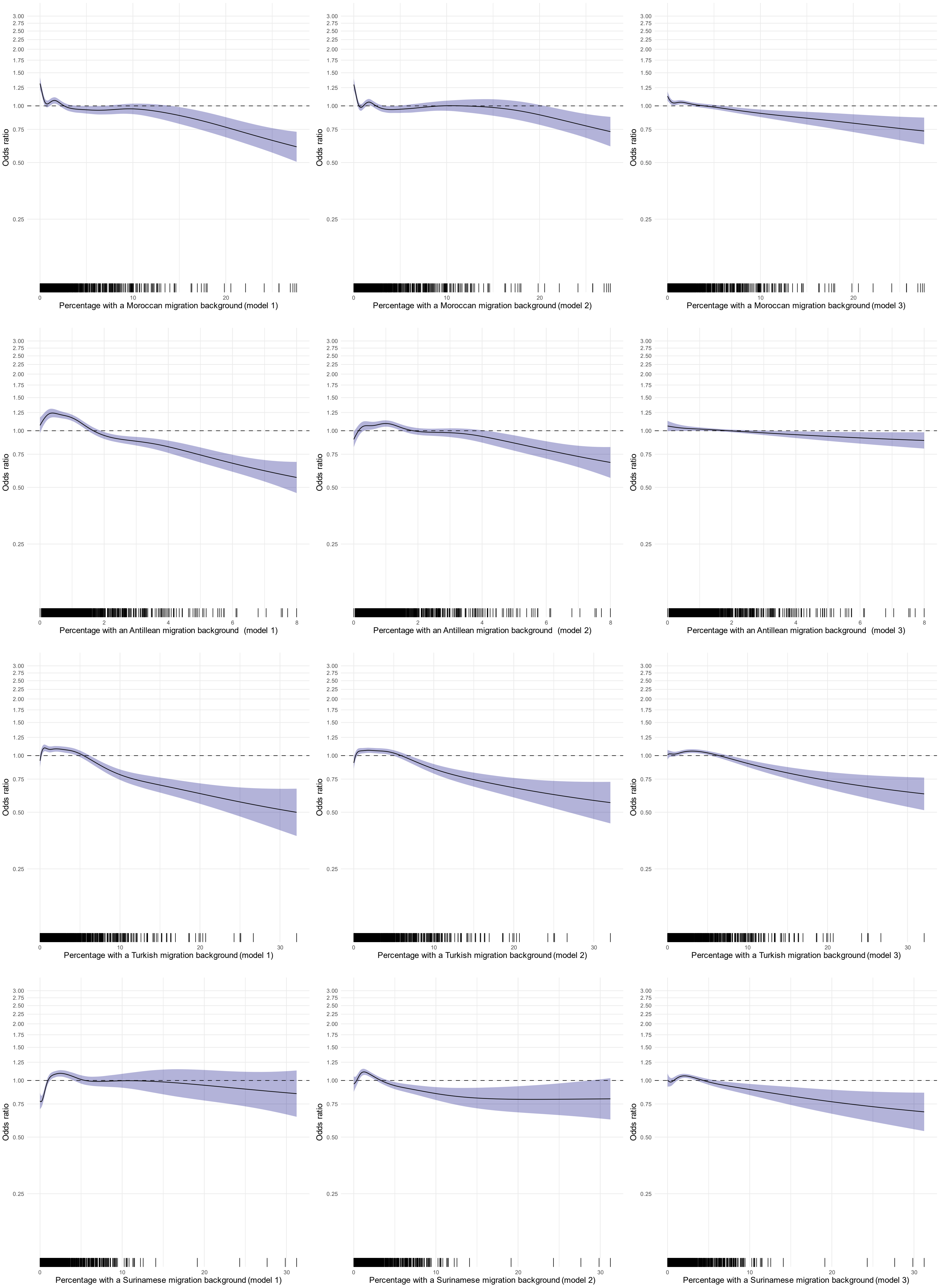
Odds ratio
3.00
2.75
2.50
2.25
2.00
1.75
1.50
1.25
1.00
0.75
0.50
0.25
Percentage with a Moroccan migration background (model 1)
Percentage with a Moroccan migration background (model 2)
Percentage with a Moroccan migration background (model 3)
Percentage with an Antillean migration background (model 1)
Percentage with an Antillean migration background (model 2)
Percentage with an Antillean migration background (model 3)
Percentage with a Turkish migration background (model 1)
Percentage with a Turkish migration background (model 2)
Percentage with a Turkish migration background (model 3)
Percentage with a Surinamese migration background (model 1)
Percentage with a Surinamese migration background (model 2)
Percentage with a Surinamese migration background (model 3)

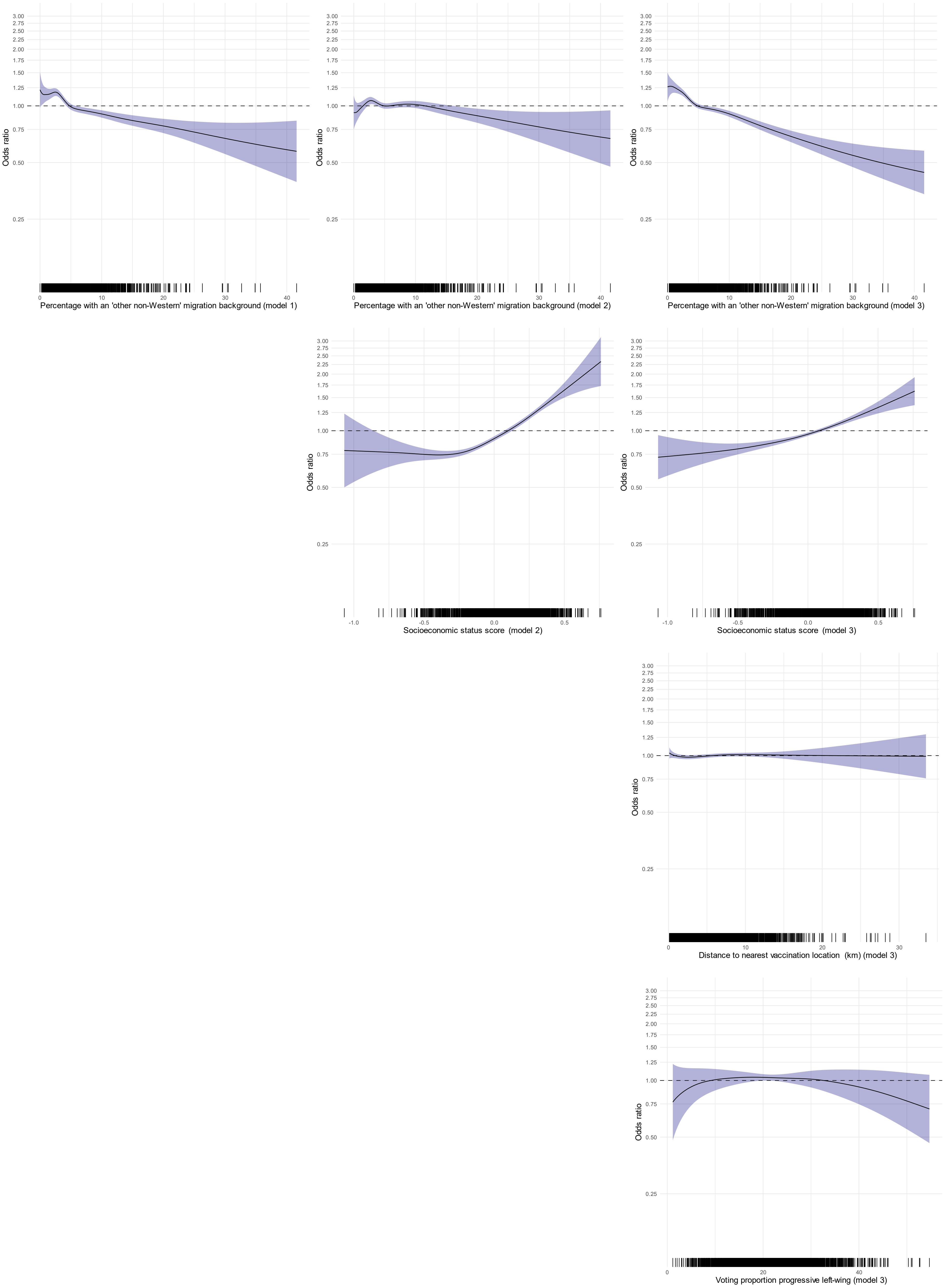
Odds ratio
3.00
2.75
2.50
2.25
2.00
1.75
1.50
1.25
1.00
0.75
0.50
0.25
0
10
20
30
40
Percentage with an 'other non-Western' migration background (model 1)
Percentage with an 'other non-Western' migration background (model 2)
Percentage with an 'other non-Western' migration background (model 3)
-1.0
-0.5
0.0
0.5
Socioeconomic status score (model 2)
Socioeconomic status score (model 3)
Distance to nearest vaccination location (km) (model 3)
Voting proportion progressive left-wing (model 3)
